# ACEI or not to ACEI: Review on using ACEI and ARBs on COVID-19 patients: Systemic review

**DOI:** 10.1101/2023.03.25.23287550

**Authors:** Ribhav Walia

## Abstract

**Purpose and motivation of this study:** Angiotensin-converting enzyme inhibitors (ACEI) and Aldosterone receptor blockers (ARBs) are one most commonly used drugs for the treatment of cardiovascular disease among other comorbidities. As multiple studies have shown that covid virus binds to the ACE2 receptor for entry into the cell. ACEI and ARBs are shown to modulate the ACE2 receptor hence, it is important to see if there are any correlations between the use of medicine and the infectivity of COVID-19. The purpose of this study is to find if the use of ACEI /ARBs can in fact increase or decrease the spread of COVID-19.

**Method:** This is a systemic review study during which all studies which helped us answer the question, of how the use of ACEI and ARBs can affect the transmission of the COVID-19 virus, were analyzed and reviewed to draw a conclusion about clinical safety and requirements of ACEI and ARBs in COVID-19 patients.

**Result:** After a complete review of all the available data very conflicting results were found. Many studies showed an increase in the transmission of COVID-19 while others showed a decreased risk of COVID-19 transmission with ACEI and ARBs use. Both results were statistically significant.

**Conclusion:** With these conflicting results the question we started with comes up again. Should we or should we not use ACEI & ARBs with covid patients? What should be the best clinical response with the use of ACEI & ARBs if it modulates transmission? To answer these, one must look not only at the statistically significant results of studies but also at the disease progression without ACEI & ARBs treatment regimes. Due to the small amount of data, there is currently no clear conclusive evidence to suggest that ACE inhibitors either increase or decrease the risk of COVID-19 transmission or the severity of the disease. Thus, more and larger studies should be developed to find a concrete answer. Until that these medicines should not be discontinued as the morbidities of cardiovascular diseases are high, and the use of ACEI and ARBs is central to the treatment.

## Introduction

In December 2019, an outbreak of SARS-CoV2 (novel coronavirus) occurred, and within a month, on January 30^th^, 2020 World Health Organisation [WHO] declared it globe pandemic [1]. It has been three years since the first outbreak of COVID-19 and as of **10 February 2023**, there have been **755**,**385**,**709 confirmed cases** of COVID-19, including **6**,**833**,**388 deaths**, reported to WHO. [2]. With the rise of covid, a need has risen for us to see how different diseases and medicine affects the pathophysiology of Covid and more importantly if there is something we can do to reduce its infectivity. Stopping the spread of covid is one of the most important steps we can take to cease the formation of new variants. Doctors and scientists began scavenging for a vaccine and an optimal treatment to decrease mortality and transmission rate. Now, we have mRNA developed by Pfizer/BioNTech, which scores high on the scale of both effectiveness and safety. Other than mRNA vaccine, inactivated vaccines (Examples include BBIBP-CorV (Sinopharm) and CoronaVac (Sinovac) and Adenovirus-vectored vaccines (example Convidecia by CanSino Biologics) are also available [3,29]. As of 30 January 2023, **13**,**168**,**935**,**724 vaccine doses** have been administered [1], yet there is a continuous rise in the death toll. In the week, **6**,**833**,**388** cumulative deaths have been reported to WHO with **755**,**385**,**709** confirmed [1]. This persistent spread of the virus has resulted in mutations in the genome of the first identified B.1.1.7 (Alpha) variant, which has resulted in the world battling not just one strain but also B.1.351 (Beta), P.1 (Gamma), B.1.617.2 (Delta), and B.1.1.529 (Omicron) variants were classified as variants of concern, which were associated with the transmission, increasing more severe disease situation (e.g., increased hospitalizations or deaths), a significant reduction in neutralization by antibodies generated during previous infection or vaccination, reduced effectiveness of treatments or vaccines, or diagnostic detection failures [4].

The rationale behind this study can be summarized as; many medicines are used including antivirals (e.g., remdesivir), antimalarials (e.g., chloroquine, hydroxychloroquine {used with azithromycin}), corticosteroids, Tocilizumab, Sarilumab, convalescent plasma, NSAIDs, and RAAS antagonists (Angiotensin-converting enzyme (ACE) inhibitors and Aldosterone receptor blockers (ARBs)) [5,29,30]. Of these, the use of ACE inhibitors and ARBs has been conflicting. A study has shown that the COVID-19 virus entry in a host cell is facilitated by ACE2 in a host cell which acts as a receptor for viral spike (S) protein [6, 26,28,30]. Another study has shown that the use of ACE inhibitors and ARBs in an animal model leads to an upregulation of ACE2 [7,15,17,24,30]. This theoretically should increase the risk of covid-19 transmission [16,19,24,30]. But some studies have shown that the use of ACE inhibitors and ARBs has soon reduced the risk of COVID-19 transmission and being symptomatically infected [8,21].

The aim of this study is to review all the evidence presented to us and draw a conclusion about whether ACEI and ARBs are beneficial in every sense or if have some back draw that should be noted by the physician before prescribing to the patient. This study answers the following question:

- Should we or should we not use ACEI & ARBs with covid patients?
- What should be the best clinical response with the use of ACEI & ARBs if it modulates transmission?

## Methodology

In this review, the Author included all types of published studies including

- systematic reviews,
- Randomized Controlled Trial,
- Books and Documents,
- Clinical Trial,
- Meta-Analysis,
- Review,
- Observational studies, and
- Clinical guidelines

Out of these Books and Documents were not found. [19]

The participants of the included studies ranged from

1. Animal subjects
2. Tissue samples that can show the regulatory effect of ACEI and ARBs on ACE2 receptor
3. Human adult Patients and/or Health adults with COVID-19 or with a risk of COVID-19 infection
4. Human adult Patients and/or Health adults with comorbidities that require treatment regimens to include ACEI and ARBs.[19]

Eligibility criterion:

All Interventions were included related to

1. the Long-term or short-term use of Angiotensin-converting enzyme inhibitors (ACEI) and Aldosterone receptor blockers (ARBs) for the treatment of a patient or a healthy adult.
2. Interaction between other drugs (e.g., antihyperglycemic drugs) and ACEI and ARBs and effect on the COVID-19 population
3. Pharmacotherapy with ACEI and ARBs was also included.
4. Drugs to treat covid-19 were included

Studies whose Outcome justifies:

1. Role of the ACE system in the infection, spread, and severity of COVID-19 was included
2. ACE2 receptor expression modulation was included
3. Effect of ACEI and ARBs on other organ systems (e.g., cardiovascular system) was included
4. Studies explaining the mechanics of Covid transmission were included

Only studies in English were included. If the subject population was pregnant, it was NOT excluded.

This study does not exclude studies on the basis of the date of publication. [19]

Reasons for exclusion were:

1. Studies published in any other language than English were excluded.
2. Any study or trial was excluded that just focused on another chronic disease (e.g., tuberculosis) that may/may not a have correlation with COVID-19.
3. study was excluded if it does not correlate the effect of the use of ACEI and ARBs with ACE 2. AND/OR Studies were excluded if it does not provide any beneficial information regarding the process of COVID-19 infectivity, severity, or spread affected by ACEI and ARBs.[19]

The author performed searches in the PubMed, and Google scholar databases on 22nd Dec 2022. Data from google scholar was excluded due to similarity with PubMed data. In addition, search the websites of the World Health Organization. WHO coronavirus disease (COVID-19) dashboard.Available at: https://covid19.who.int Accessed 13 Feb 2023. We hand-searched reference lists of included studies and previously published reviews.[19]

The author searched PubMed on 22^nd^ Dec 2022. The database coverage does not exclude studies on the basis of the date of publication. But most of the relevant results for this study were after 2020 (after the outbreak of the virus). [19]

The advanced search on PubMed was made using the following search terms:

1. “Angiotensin-Converting Enzyme Inhibitors”[Mesh]
2. OR “Angiotensin-Converting Enzyme inhibitor*” [tw]
3. “Angiotensin Receptor Antagonists”[Mesh]
4. OR “Angiotensin Receptor blocker*”[tw]
5. “COVID-19”[Mesh]
6. OR “SARS-CoV-2”[Mesh]
7. OR COVID-19 [tw]
8. “Disease Transmission, Infectious”[Mesh]
9. OR Transmission [tw]
10. “Angiotensin-Converting Enzyme 2”[Mesh]
11. OR “Angiotensin-Converting Enzyme 2 receptor*”
12. “COVID-19”[Mesh],
13. “SARS-CoV-2”[Mesh]
14. “Disease Transmission, Infectious”[Mesh]

Based on the search, we identified 25 completed studies out of which we found 16 of them eligible for inclusion in our review as they meet the eligibility criteria. Further, we hand-searched reference lists of included studies and previously published reviews and added 12 more studies that meet our eligibility criteria. The author independently screened titles and abstracts of all retrieved articles. In case of confusion, consensus on articles to screen full-text was reached.[19]

**Table 1:** The studies obtained initially after the advanced search on PubMed can be presented on the bases on the year of publication as follows: [19]

| Search query: (((("Angiotensin-Converting Enzyme Inhibitors"[Mesh] OR "Angiotensin-Converting Enzyme inhibitor*" [tw]) OR ("Angiotensin Receptor Antagonists"[Mesh] OR "Angiotensin Receptor blocker*" [tw])) AND ("COVID-19"[Mesh] OR "SARS-CoV-2"[Mesh] OR "COVID-19*" [tw])) AND ("Disease Transmission, Infectious"[Mesh] OR Transmission [tw])) |  |
| --- | --- |
| Year | Count |
| 2022 | 3 |
| 2021 | 8 |
| 2020 | 15 |

**Table 2:** The advanced search on PubMed using search terms initially presented below:[19]

| Search number | Query | Sort By | Filters | Search Details | Results | Time |
| --- | --- | --- | --- | --- | --- | --- |
| 5 | ((("Angiotensin Receptor Antagonists"[Mesh] OR "Angiotensin Receptor blocker*"[tw]) OR ("Angiotensin-Converting Enzyme Inhibitors"[Mesh] OR "Angiotensin-Converting Enzyme inhibitor*" [tw])) AND ("COVID-19"[Mesh] OR "SARS-CoV-2"[Mesh] OR COVID-19 [tw])) AND ("Disease Transmission, Infectious"[Mesh] OR Transmission [tw])) |  |  | ("Angiotensin Receptor Antagonists"[MeSH Terms] OR "angiotensin receptor blocker*"[Text Word] OR ("Angiotensin-Converting Enzyme Inhibitors"[MeSH Terms] OR "angiotensin converting enzyme inhibitor*"[Text Word])) AND ("COVID-19"[MeSH Terms] OR "SARS-CoV-2"[MeSH Terms] OR "COVID-19"[Text Word]) AND ("disease transmission, infectious"[MeSH Terms] OR "Transmission"[Text Word]) | 26 | 13:44:47 PM |
| 4 | "Disease Transmission, Infectious"[Mesh] OR Transmission [tw] |  |  | "disease transmission, infectious"[MeSH Terms] OR "Transmission"[Text Word] | 6,17,753 | 13:42:47 PM |
| 3 | "COVID-19"[Mesh] OR "SARS-CoV-2"[Mesh] OR COVID-19 [tw] |  |  | "COVID-19"[MeSH Terms] OR "SARS-CoV-2"[MeSH Terms] OR "COVID-19"[Text Word] | 2,72,330 | 13:14:52 PM |
| 2 | "Angiotensin Receptor Antagonists"[Mesh] OR "Angiotensin Receptor blocker*"[tw] |  |  | "Angiotensin Receptor Antagonists"[MeSH Terms] OR "angiotensin receptor blocker*"[Text Word] | 23,971 | 13:14:26 PM |
| 1 | "Angiotensin-Converting Enzyme Inhibitors"[Mesh] OR "Angiotensin-Converting Enzyme inhibitor*" [tw] |  |  | "Angiotensin-Converting Enzyme Inhibitors"[MeSH Terms] OR "angiotensin converting enzyme inhibitor*"[Text Word] | 45,025 | 13:14:05 PM |

From these 26 studies, only 18 were selected based on the eligibility criteria described above. The result obtained was screened via title and abstract and its relevance to the topic was discussed with mentors. After the initial screening data from each study were extracted onto a spreadsheet and then further refined to use. This process was repeated for all 18 studies obtained from PubMed and also the other 11 studies that were hand-picked from the list of included studies. If these studies 2 were just abstract and data was extracted from just the abstract. one study only had incomplete data available publicly and only that was used. 1 study was a clinical trial that is still yet to provide results. The initial screening If the study’s title and abstract were not providing sufficient information to either include or exclude the study on the bases of set eligibility criteria the full text was screened and then the decision was made. [19]

All studies were summarized in a tabular format. This table comprised summaries of the estimated intervention effect and the number of participants and studies for each primary outcome. The table was reviewed and a rational decision was made to formulate a result and conclusion. [19]

Using a flow diagram, the results of the search and selection process, from the number of records identified in the search to the number of studies included in the review, are displayed below:

In our study we excluded 7 studies [19] :

- 1 study (Kreutz R, Abd El-Hady Algharably E, Ganten D, Messerli F. Renin-Angiotensin-System (RAS) und COVID-19 – Zur Verordnung von RAS-Blockern [Renin-Angiotensin-System (RAS) and COVID-19 - On The Prescription of RAS Blockers]. Dtsch Med Wochenschr. 2020 May;145(10):682-686. German. doi: 10.1055/a-1152-3469. Epub 2020 Apr 22. PMID: 32323279; PMCID: <u>PMC7295302.</u> https://pubmed.ncbi.nlm.nih.gov/32323279/) was identified to be in the German language and was excluded.
- 2 studies (Hussain A, Bhowmik B, do Vale Moreira NC. COVID-19 and diabetes: Knowledge in progress. Diabetes Res Clin Pract. 2020 Apr;162:108142. doi: 10.1016/j.diabres.2020.108142. Epub 2020 Apr 9. PMID: 32278764; PMCID: PMC7144611. https://pubmed.ncbi.nlm.nih.gov/32278764/) and (Gül Ş, Akalın Karaca ES, Özgün Niksarlıoğlu EY, Çınarka H, Uysal MA. Coexistence of tuberculosis and COVID-19 pneumonia: A presentation of 16 patients from Turkey with their clinical features. Tuberk Toraks. 2022 Mar;70(1):8-14. English. doi: 10.5578/tt.20229902. PMID: 35362300. https://pubmed.ncbi.nlm.nih.gov/35362300/) was excluded because they just focused on another chronic disease (e.g., tuberculosis) that may/may not a have correlation with COVID-19. (Gül Ş, Akalın Karaca ES, Özgün Niksarlıoğlu EY, Çınarka H, Uysal MA. Coexistence of tuberculosis and COVID-19 pneumonia: A presentation of 16 patients from Turkey with their clinical features. Tuberk Toraks. 2022 Mar;70(1):8-14. English. doi: 10.5578/tt.20229902. PMID: 35362300. https://pubmed.ncbi.nlm.nih.gov/35362300/) was also excluded because it do not correlate the effect of the use of ACEI and ARBs with ACE 2 and/or Studies were excluded if it does not provide any beneficial information regarding the process of COVID-19 infectivity, severity, or spread affected by ACEI and ARBs.
- 5 studies (Kreutz R, Abd El-Hady Algharably E, Ganten D, Messerli F. Renin-Angiotensin-System (RAS) und COVID-19 – Zur Verordnung von RAS-Blockern [Renin-Angiotensin-System (RAS) and COVID-19 - On The Prescription of RAS Blockers]. Dtsch Med Wochenschr. 2020 May;145(10):682-686. German. doi: 10.1055/a-1152-3469. Epub 2020 Apr 22. PMID: 32323279; PMCID: PMC7295302. https://pubmed.ncbi.nlm.nih.gov/32323279/), (Schellack N, Coetzee M, Schellack G, Gijzelaar M, Hassim Z, Milne M, Bronkhorst E, Padayachee N, Singh N, Kolman S, Gray AL. COVID-19: Guidelines for pharmacists in South Africa. S Afr J Infect Dis. 2020 Jun 10;35(1):206. doi: 10.4102/sajid.v35i1.206. PMID: 34192121; PMCID: PMC7577345. https://pubmed.ncbi.nlm.nih.gov/34192121/), (Axiotakis LG Jr, Youngerman BE, Casals RK, Cooke TS, Winston GM, Chang CL, Boyett DM, Lalwani AK, McKhann GM. Risk of Acquiring Perioperative COVID-19 During the Initial Pandemic Peak: A Retrospective Cohort Study. Ann Surg. 2021 Jan 1;273(1):41-48. doi: 10.1097/SLA.0000000000004586. PMID: 33156061; PMCID: PMC7737880. https://pubmed.ncbi.nlm.nih.gov/33156061/), (Skayem C, Ayoub N. Carvedilol and COVID-19: A Potential Role in Reducing Infectivity and Infection Severity of SARS-CoV-2. Am J Med Sci. 2020 Sep;360(3):300. doi: 10.1016/j.amjms.2020.05.030. Epub 2020 May 28. PMID: 32631576; PMCID: PMC7833103. https://pubmed.ncbi.nlm.nih.gov/32631576/) and (Ruocco G, Feola M, Palazzuoli A. Hypertension prevalence in human coronavirus disease: the role of ACE system in infection spread and severity. Int J Infect Dis. 2020 Jun;95:373-375. doi: 10.1016/j.ijid.2020.04.058. Epub 2020 Apr 24. PMID: 32335337; PMCID: PMC7180155. https://pubmed.ncbi.nlm.nih.gov/32335337/) were excluded because they do not correlate the effect of the use of ACEI and ARBs with ACE 2 and/or Studies were excluded if it does not provide any beneficial information regarding the process of COVID-19 infectivity, severity, or spread affected by ACEI and ARBs.

Bias could have been arisen due to any of the following factor:

1. bias arising from the data review process;
2. bias due to missing / incomplete data
3. bias due to only a single author being involved in the completion of the study (paper only reviewed by one mentor)
4. bias due to missing outcome data;
5. bias in the measurement of the outcome; and
6. bias in the selection of the reported result. [19]

Only one author was involved in the review process independently and independently applied the tool to each included study, and recorded supporting information and justifications for judgments of risk of bias for each domain (low; high; some concerns). Any discrepancies in judgments of risk of bias or justifications for judgments were resolved by discussing with mentors.[19]

Table 3: The table below contains

- The characteristics of each study included in this systemic review.
- Characteristics of the included clinical trials.
- Risk of bias where represented with the following symbols: ↑= low, ↑↑= high, ↑↑↑= some concerns
- Results of individual studies
- Results of syntheses
- Outcome
- Certainty of evidence: in the table “+” represents that the author is certain about the presence of evidence in the study.

## Discussion

### Entry of COVID-19

To understand how COVID-19 can be affected by ACE inhibitors and ARBs we must understand the entry of COVID-19 into the human cell. Further understanding the cellular factors that are used by SARS-CoV-2 for entry might provide insights into viral transmission and help discover the therapeutic targets [6,26,27,28]. The virus has a structure that includes the spike protein on the surface. This spike protein binds to the receptor on the human cell called ACE2, which is found in the various tissue inside the body, including the lungs, heart, kidneys, and intestine. [23, 27,30]

The spike (S) protein of SARS-CoV2 has two subunits; S1 and S2[6, 25, 27,28,30]. Entry of covid virus depends on subunit S1 binding to the cellular receptor, angiotensin-converting enzyme 2(ACE2), on the host cell. As S1 attaches to the ACE2; cellular protease, transmembrane protease serine-type 2 (TMPRSS2) TMPRSS2, and cleaves S protein at the S1/S2 site and S2’ [6, 9, 25, 27,30]. This process is called priming. Other host cell proteases, such as cathepsin B/L, can also achieve it. Cathepsin B/L is not essential for viral spread and pathogenesis, unlike the priming by TMPRSS2[6, 9, 16, 25]. Priming of S1 allows the fusion of the virus into the cell membrane which is mediated by the S2 subunit [6,16,25,28,30].

These structural changes in the spike protein of the virus allow the virus to fuse with the human cell’s membrane. This fusion allows the virus to release its genetic material, which is RNA, into the human cell. Once inside the cell, the viral RNA is used by the cell’s machinery to produce new viral proteins and RNA, which assemble into new virus particles. Which further infects the new cell. [25,27,28,29]

### The effect of ACE inhibitors on ACE 2 receptors

ACE inhibitors and ARBs are two drugs commonly used to treat high blood pressure and heart failure. The concern about these drugs and the upregulation of the ACE2 receptor arose because ACE2 is the receptor that the SARS-CoV-2 virus uses to enter human cells.[15,26] Classically, Due to a fall in blood pressure or blood volume, Renin converts a protein called angiotensinogen, which is produced by the liver, into angiotensin I. Angiotensin I travels to the lungs, where an enzyme called angiotensin-converting enzyme (ACE) converts it into angiotensin II. Angiotensin II causes blood vessels to constrict, increasing blood pressure, and stimulating the release of the hormone aldosterone from the adrenal glands. [10,11,15, 23]

The function of the ACE2 enzyme in the RAAS is to act as a counter-regulatory mechanism to the actions of the angiotensin-converting enzyme (ACE) [15,23]. ACE2 is an enzyme that is located on the surface of cells in various organs and tissues, including the lungs, heart, kidneys, and gastrointestinal tract. It converts angiotensin II (Ang II), a potent vasoconstrictor and pro-inflammatory molecule, into angiotensin-(1-7) [Ang-(1-7)], a vasodilator and anti-inflammatory molecule [17]. This conversion by ACE2 results in a decrease in the levels of Ang II and an increase in the levels of Ang-(1-7), leading to a decrease in blood pressure and inflammation. [11,12,13,17,23]

ACE inhibitors work by blocking the activity of an enzyme called angiotensin-converting enzyme (ACE), which is responsible for converting angiotensin I to angiotensin II. By blocking the action of ACE, ACE inhibitors reduce the levels of angiotensin II in the body. ARBs work by blocking the action of angiotensin II at its receptor sites, preventing its effects on blood vessels and other organs [15,17,23].

The effects of ace and arbs are depicted in the image below

**Fig. 1.**
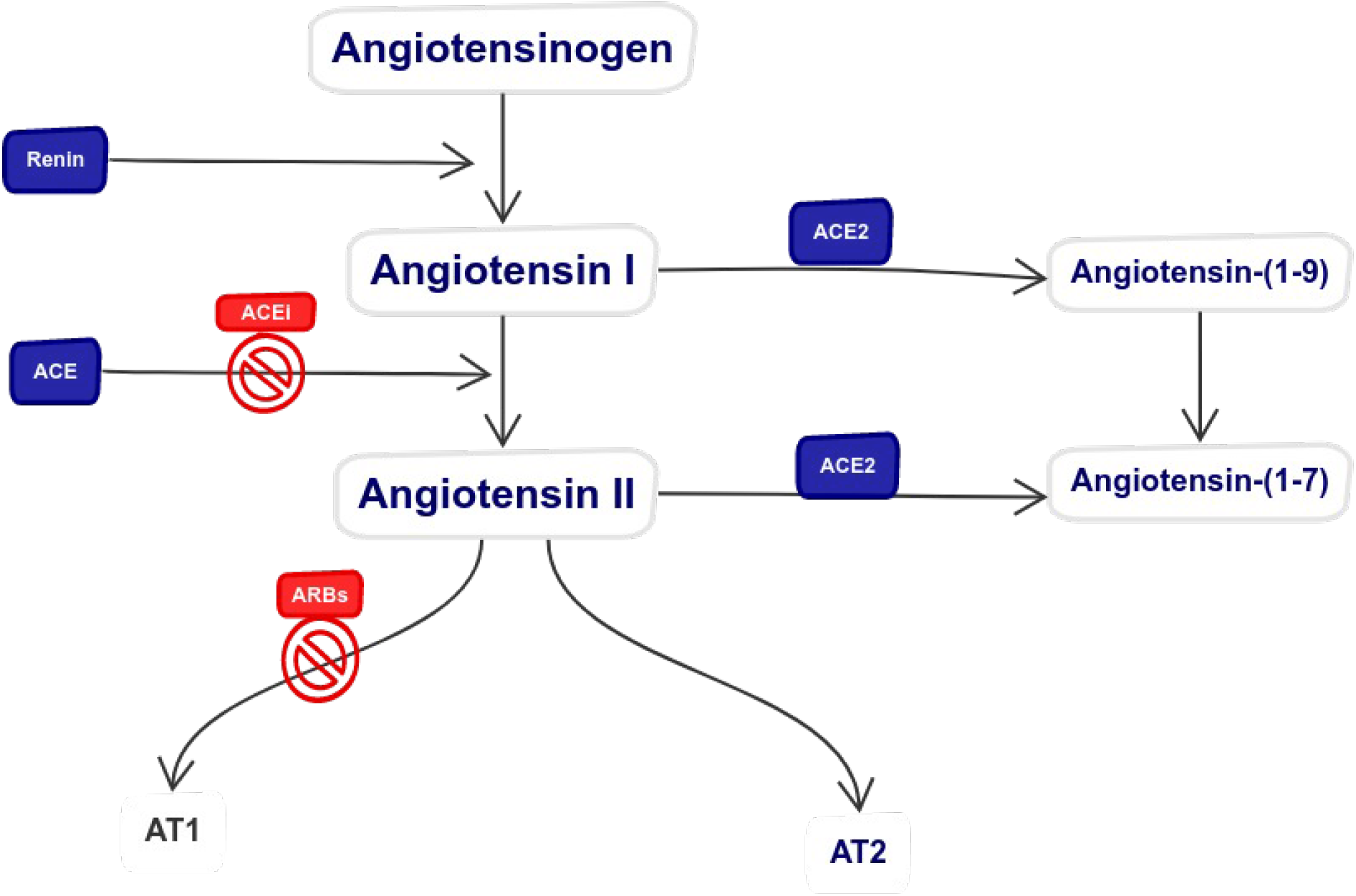
The RAS pathway shows the classical and protective arms as well as the various receptors and their downstream effects. RAS: Renin Angiotensin System. ACEI: Angiotensin Converting Enzyme inhibitor. ARB: Angiotensin Receptor Blocker. AT1: Angiotensin 1 receptor. AT2: Angiotensin 2 receptor.

Both ACE inhibitors and ARBs have been shown to increase the expression of the ACE2 receptor in the body [5,15,17,23]. Thus, RAAS should, theoretically, influence ACE2 expression and/or activity and may also influence the susceptibility to infection, the severity the of infection, and potential complications, for those who are once infected [17,23].

## RESULT

It is important to note that while there is some evidence to suggest that ACEI and ARBs may increase ACE2 expression, the overall effect of these medications on COVID-19 outcomes is still under investigation and remains controversial. Some studies have suggested that ACEI and ARBs may have a protective effect against severe COVID-19, while others have found no significant association or even a potentially harmful effect. According to some studies, ACEI and ARBs are known to increase the ACE2 levels. Studies have suggested the risk of developing COVID-19 with the administration of ACEI and ARBs as they can overproduce the circulating ACE2 transcripts in the cells [15],[16].

A study reported that the drug captopril (ACEI family drug) can regulate ACE2 overexpression while downregulating the angiotensin II-dependent AT1 receptor downstream signalling and RANKL expression in osteoporotic rats. ACE2 expression was considerably upregulated in the study in rats that were given captopril. Thus, captopril (ACEI) demonstrated a clinical role in the upregulation of the ACE2-dependent Mas receptor signaling cascade in restoring bone metabolism [17]. Another study showed that Enalapril increases ACE2 expression in the kidney tissues, but no significant fold change was observed in TMPRSS2 mRNA expression [17], [18].

Another experiment study mice comprised of a drug treatment group with lisinopril, losartan, lisinopril, and losartan combined, or vehicle over a course of 42 days. The study showed increased expression of ACE2 mRNA while the optimum reduction in angiotensin II (plasma) expression in the group treated with ACEI(lisinopril) and ARBs (losartan). The important conclusion that can be derived from this study is two conflicting results as there was an increased ACE2 protein in the lung and small intestine. In contrast, the combination of lisinopril with losartan prevented the lisinopril-induced increase in tissue ACE2 levels. And so, the expression of ACE2 is different in different tissue [31].

Another retrospective study enrolling 452 people living in retirement homes (PLRNH) suggests that a significant difference between COVID-19-infected and not-infected. Of those receiving chronic treatment with Angiotensin II receptor blockers (ARBs) (18.6% vs. 9.5%, p=0.012). On the contrary, there was no difference in the proportion of those receiving ACE inhibitors (ACE-I) (21.2% vs. 23.6%, p=0.562). Receiving ARBs as a chronic treatment medication in close communities act as an independent predictor of infection risk [OR 1.95 (95% CI 1.03-3.72) p=0.041] [24].

Although some researchers have suggested their concerns regarding the association of ACEI and ARBs with the ACE2 upregulation in COVID-19-infected comorbidities and have suggested switching towards an alternative pharmacological agent for therapeutic intervention [15].

Another study from Stanford Sinus Centre, National Taiwan University Hospital (NTUH), and China Medical University Hospital (CMUH) in Taiwan showed the use of ACEI or ARBs does not increase ACE2 expression in the upper respiratory cilia, and therefore patients taking ACEI or ARBs are likely at no greater risk of SARS-CoV-2 transmission than individuals not on these medications[20]. This was a cohort study in which patients were identified within the sinonasal tissue bank. They were grouped as those who have been taking either ACEI or ARBs for at least six continuous months prior to sinonasal surgery and those who have never taken ACEI/ARBs. the control was matched for their controls matched for ages, sex, and smoking status. The groups were then compared for their ACE2 expression. The study reports that ciliary ACE2 expression is slightly, but statistically significantly, decreased in patients taking ACEI compared to matched controls, whereas ACE2 expression was not significantly different in patients taking ARBs compared to controls. The study also produced a Subgroup analysis comparing ACEI and ARBs treatment groups to controls of hypertension, similar age, sex, or smoking status revealing a similar trend of non-significant but lower ACE2 expression in the ACEI and ARB group[20]. Thus, this study, unambiguously indicates that the use of ACEI or ARBs does not increase ACE2 expression in the upper respiratory cilia [20].

Another study, a phase 2B, prospective, randomized, double-blind, allocation-concealed, placebo-controlled, multicentre study evaluated the effect of once-daily ramipril (2.5 mg orally) versus placebo for 14 days. And found ACEI, ramipril, to decrease ICU admission, the requirement for mechanical ventilation, and death. The study concluded ACEI drugs have longstanding safety data and a putative mechanistic hypothesis for benefit in COVID-19 [21].

One study included 1,499 of the 9,101 household contacts who were taking an ACEI or an ARBs. This study showed that the Probability of COVID-19 diagnosis was slightly higher among ACEI/ARB users. However, ACEI/ARB users were older and when the cohort was adjusted for age, gender, race/ethnicity, English proficiency, comorbid conditions, time period, including patients with documented hypertension, diabetes, or cardiovascular disease, as well as when including other medications in the models, use of ACEI/ARBs remained associated with a decreased risk of COVID-19 infection in propensity score analyses with a predicted probability of COVID infection 18.6% in ACEI/ARB users vs. 24.5% in non-users, p = 0.03 [8].

## Conclusion

It is important to note that while there is some evidence to suggest that ACEI and ARBs may increase ACE2 expression, the overall effect of these medications on COVID-19 outcomes is still under investigation and remains controversial. Some studies have suggested that ACEI and ARBs may have a protective effect against severe COVID-19, while others have found no significant association or even a potentially harmful effect. Because of the complex relationship between ACE inhibitors and COVID-19, a review article concludes that although theoretically, RAS inhibition may lead to increased susceptibility, disease severity, and mortality in COVID-19 patients receiving ACEI or ARBs, there has been no clinical evidence based on observational studies supporting such an outcome in patients with COVID-19 infection. However, properly designed will be needed to further confirm or refute current evidence. Until such evidence becomes available, guideline-directed therapies should be followed as currently there is no need to withdraw RAS blockers in stable patients during the COVID-19 pandemic [14], [22].

In fact, some studies have suggested that ACE inhibitors and similar drugs may actually have a protective effect against severe COVID-19, potentially by reducing inflammation or by mitigating the effects of hypertension, which is a known risk factor for severe disease.

To conclude, there is still a lack of sufficient amount of data to support whether the withdrawal of ACE and ARBs medication provides any benefits or risks to a patient. But sufficient data is available on the effects of lack of ACEI and ARBs in the medical regime of a patient suffering from comorbidities that required them to have ACEI and ARBs.

There were initial concerns that the use of ACE inhibitors might increase the risk of COVID-19 transmission or the severity of the disease, as these drugs increase the expression of ACE2 in the body, potentially providing more entry points for the virus. But ACE inhibitors are a class of drugs that are commonly used to treat high blood pressure and heart failure. The morbidity of these diseases is high without treatment. [22,24]

## Supporting information

Methodology

Flow chart 1

## Data Availability

All data produced are available online at

https://www.ncbi.nlm.nih.gov/pmc/articles/PMC7068984/

https://www.ncbi.nlm.nih.gov/pmc/articles/PMC9027683/

https://www.ncbi.nlm.nih.gov/pmc/articles/PMC9126103/

https://www.sciencedirect.com/science/article/pii/S0735675720302631

https://www.ncbi.nlm.nih.gov/pmc/articles/PMC7102627/

https://www.ahajournals.org/doi/10.1161/CIRCULATIONAHA.104.510461?url_ver=Z39.88-2003&rfr_id=ori:rid:crossref.org&rfr_dat=cr_pub%20%200pubmed

https://www.ncbi.nlm.nih.gov/pmc/articles/PMC7924745/

https://www.ncbi.nlm.nih.gov/pmc/articles/PMC7665323/

https://doi.org/10.1038/s41569-019-0244-8

## Abbreviations

RAAS: renin angiotensin aldosterone system
RAS: Renin Angiotensin System
ACE: Angiotensin-converting enzyme
ACEI: Angiotensin-converting enzyme inhibitor
ARBs: Aldosterone receptor blockers
WHO: World Health Organisation
TMPRSS2: Transmembrane protease serine-type 2
SARS-CoV2: Severe Acute Respiratory Syndrome Coronavirus 2.
COVID-19: Coronavirus disease 2019
Ang: Angiotensin

---

**Flow chart 1.**
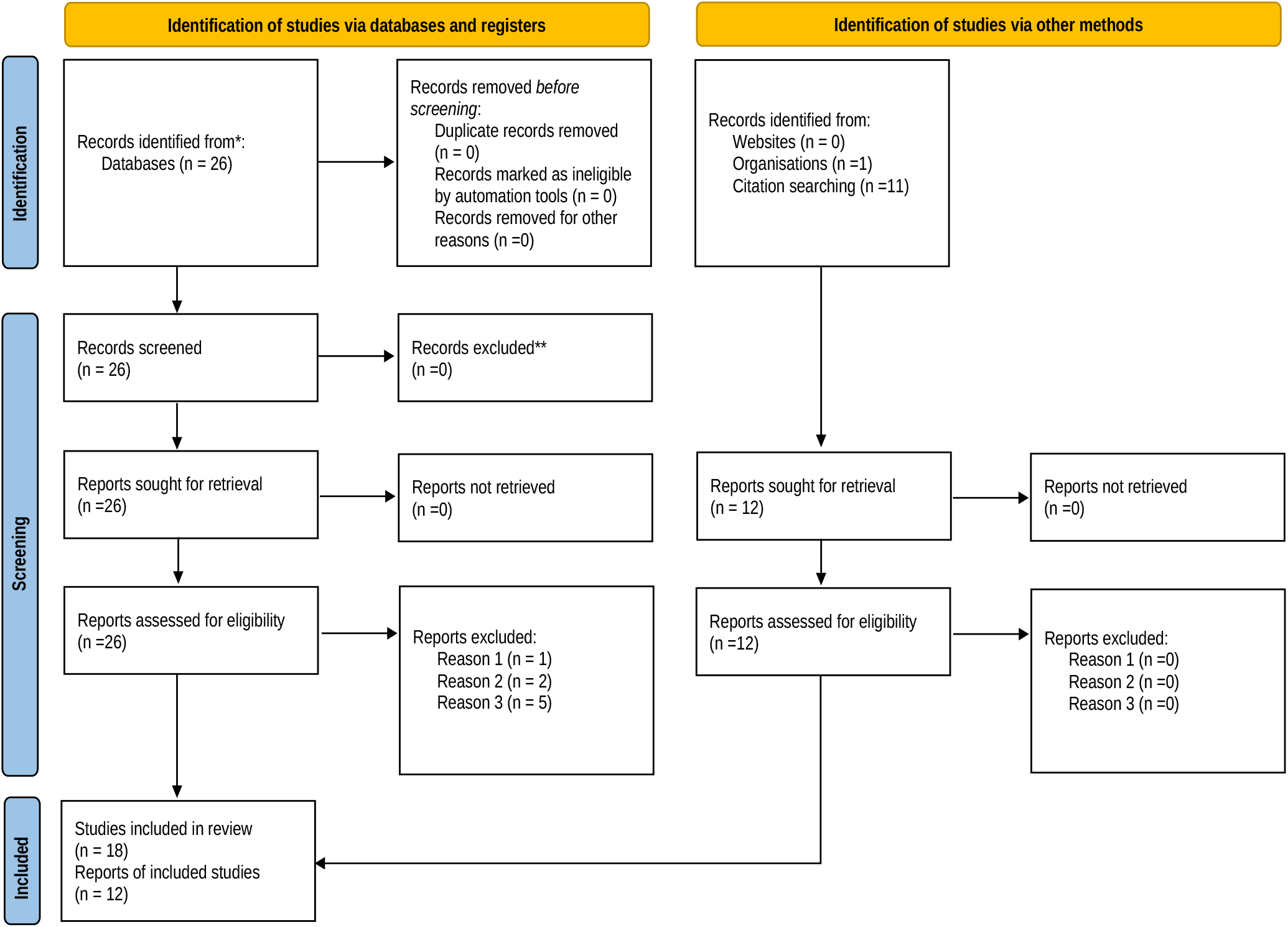
PRISMA 2020 flow diagram updated *Consider, if feasible to do so, reporting the number of records identified from each database or register searched (rather than the total number across all databases/registers). **If automation tools were used, indicate how many records were excluded by a human and how many were excluded by automation tools. *From:* Page MJ, McKenzie JE, Bossuyt PM, Boutron I, Hoffmann TC, Mulrow CD, et al. The PRISMA 2020 statement: an updated guideline for reporting systematic reviews. BMJ 2021;372:n71. doi: 10.1136/bmj.n71. For more information, visit: http://www.prisma-statement.org/

