## Supplementary material for "ACEI or not to ACEI: Review on using ACEI and ARBs on COVID-19 patients: Systemic review": Flow chart 1

**Identification of studies via other methods**

**Identification of studies via databases and registers**

Records identified from:

Websites (n = 0)

Organisations (n =1)

Citation searching (n =11)

Records removed *before screening*:

Duplicate records removed (n = 0)

Records marked as ineligible by automation tools (n = 0)

Records removed for other reasons (n =0)

Records identified from*:

Databases (n = 26)

**Identification**

Records screened

(n = 26)

Records excluded**

(n =0)

Reports not retrieved

(n =0)

Reports sought for retrieval

(n = 12)

Reports sought for retrieval

(n =26)

Reports not retrieved

(n =0)

**Screening**

Reports assessed for eligibility

(n =12)

Reports excluded:

Reason 1 (n =0)

Reason 2 (n =0)

Reason 3 (n =0)

Reports assessed for eligibility

(n =26)

Reports excluded:

Reason 1 (n = 1)

Reason 2 (n = 2)

Reason 3 (n = 5)

Studies included in review

(n = 18)

Reports of included studies

(n = 12)

**Included**

*Consider, if feasible to do so, reporting the number of records identified from each database or register searched (rather than the total number across all databases/registers).

**If automation tools were used, indicate how many records were excluded by a human and how many were excluded by automation tools.

*From:*  Page MJ, McKenzie JE, Bossuyt PM, Boutron I, Hoffmann TC, Mulrow CD, et al. The PRISMA 2020 statement: an updated guideline for reporting systematic reviews. BMJ 2021;372:n71. doi: 10.1136/bmj.n71. For more information, visit: <http://www.prisma-statement.org/>
